# Invisible Text Injection: The Trojan Horse of AI-Assisted Medical Peer Review

**DOI:** 10.1101/2025.07.24.25332148

**Authors:** Byungjin Choi, Tae Joon Jun, Jong Won Sung, Ilwoo Park, Jeong-Moo Lee, Soo Ick Cho, Hyung Jun Park, Ro Woon Lee, Jungyo Suh

**Author notes:** Corresponding author: Jungyo Suh, MD, Department of Urology, Asan Medical Centre, Ulsan University College of Medicine 88, Olympic-ro 43-gil, Songpa-gu, 05505, Seoul, Republic of Korea.

## Abstract

**Key Points:** *Question:* Are large language models robust against adversarial attacks in medical peer review?

*Findings:* In this factorial experimental study, invisible text injection attacks significantly increased review scores and raised manuscript acceptance rates from 0% to nearly 100%, while also significantly impairing the ability of large language models to detect scientific flaws.

*Meaning:* Enhanced safeguards and human oversight are essential prerequisite for using large language models in medical peer review.

**Importance:** Large language models (LLMs) are increasingly considered for medical peer review. However, their vulnerability to adversarial attacks and ability to detect scientific flaws remain poorly understood.

**Objective:** Evaluate LLMs’ ability to identify scientific flaws in peer review and their robustness against invisible text injection (ITI).

**Design, Setting, and Participants:** This factorial experimental study was conducted in May 2025 using a 3 LLMs × 3 prompt strategies × 4 manuscript variants x 2 with/without ITI design. We used three commercial LLMs (Anthropic, Google, OpenAI). The four manuscript variants either contained no flaws (control) or included scientific flaws in the methodology, results, or discussion section, respectively. Three prompt strategies were evaluated: neutral peer review, strict guidelines emphasizing objectivity, and explicit rejection.

**Interventions:** ITI involved inserting concealed instructions using white text on white background, directing LLMs to review with positive evaluations and “accept without revision” recommendations.

**Main Outcomes and Measures:** Primary outcomes were review scores (1-5 scale) and acceptance rates under neutral prompts. Secondary outcomes were review scores, acceptance rates under strict and explicit reject prompts. We investigated flaw detection capability using liberal (detect any flaw) and stringent (detect all flaw) criteria. We calculated mean score differences by models and prompt types and used t-test and Fisher’s exact test for calculating P-value.

**Results:** ITI caused significant score inflation under neutral prompts. Score differences for Anthropic, Google and OpenAI were 1.0 (P<.001), 2.5 (P<.001) and 1.7 (P<.001). Acceptance rates increased from 0% to 99.2%-100% across all providers (P<.001). Score differences were still statistically significant under strict prompting. Score differences were not significant under explicit rejection prompting, but flaw detection rate was still impaired. Using liberal detection criteria, results section flaw detection rate was significantly compromised with ITI, particularly in Google (88.9% to 47.8%, P<.001). Stringent criteria revealed methodology detection falling from 56.3% to 25.6% (P<.001) and overall detection dropping from 18.9% to 8.5% (P<.001).

**Conclusions and Relevance:** ITI can significantly alter the evaluation of medical studies by LLMs, and mitigation at the prompt level is insufficient. Enhanced safeguards and human oversight are essential prerequisites for the application of LLMs in medical publishing.

## Introduction

The advancement of medical science and its translation into clinical practice depend on the integrity of peer review.^1^ This gatekeeping process upholds the standards of medical literature by scrutinizing manuscript validity, significance, and originality, thereby directly influencing patient care and public health.^2,3^ However, peer review is labor-intensive, often relying on the goodwill of researchers without direct incentives. ^2–4^ This systemic pressure, coupled with exponentially increasing scholarly output, has prompted active exploration of Large Language Models (LLMs) to enhance review efficiency.^5–9^ Consequently, LLMs are increasingly likely to encounter manuscript data, regardless of official journal policies prohibiting their use.

Despite growing interest in LLM-assisted peer review, fundamental questions about their reliability and safety remain. ^5^ Recent evidence suggests that LLMs are susceptible to adversarial attacks such as invisible text injection (ITI), where malicious, imperceptible text is embedded within manuscripts.^9–11^ Such attacks can force predetermined conclusions by embedding hidden instructions readable only to LLMs. While prior studies have demonstrated LLM vulnerabilities in general contexts^10,11^, no research has systematically evaluated how adversarial manipulation affects medical research paper peer review quality. Invisible text injection (ITI) can be a threat to the medical science community by manipulating LLMs during peer review, compelling them to produce inappropriately favorable evaluations irrespective of manuscript quality. This theoretical concern has now become a reality, as a case was recently reported where researchers from world-leading universities embedded hidden prompts in papers to induce positive reviews.^12^ The probability of the same occurring in the medical field is high.

In this study, we aim to systematically examines whether ITI attacks can manipulate LLM evaluations of scientific manuscripts, using three leading commercial models (Anthropic Claude, Google Gemini, and OpenAI GPT). As a secondary objective, we assess these models’ baseline ability to detect substantive scientific flaws across different manuscript sections: methodology, results interpretation, and discussion.

## Methods

### Study Design and Setting

This controlled experimental study was conducted in May 2025 to evaluate LLM performance and vulnerability in simulated peer review scenarios using ITI across multiple prompting conditions. The experiment employed a fully crossed factorial design, 3 LLM providers × 3 prompt types × 4 manuscript variants x 2 with/without ITI × 30 replications, yielding 2160 total API calls. Each experimental condition was replicated 30 times to ensure statistical power for detecting meaningful differences. Replications were conducted sequentially with systematic rotation through all experimental conditions to control for temporal effects and API performance variations. Technical safeguards included automated error handling with retry mechanisms for failed API calls (maximum 3 attempts with exponential backoff), interim data preservation every 10 completed responses to prevent data loss, and comprehensive logging of all API response metadata including timestamps, model versions, and response latencies. Details of experimental design and procedure described in **Figure 1**

**Figure 1.**
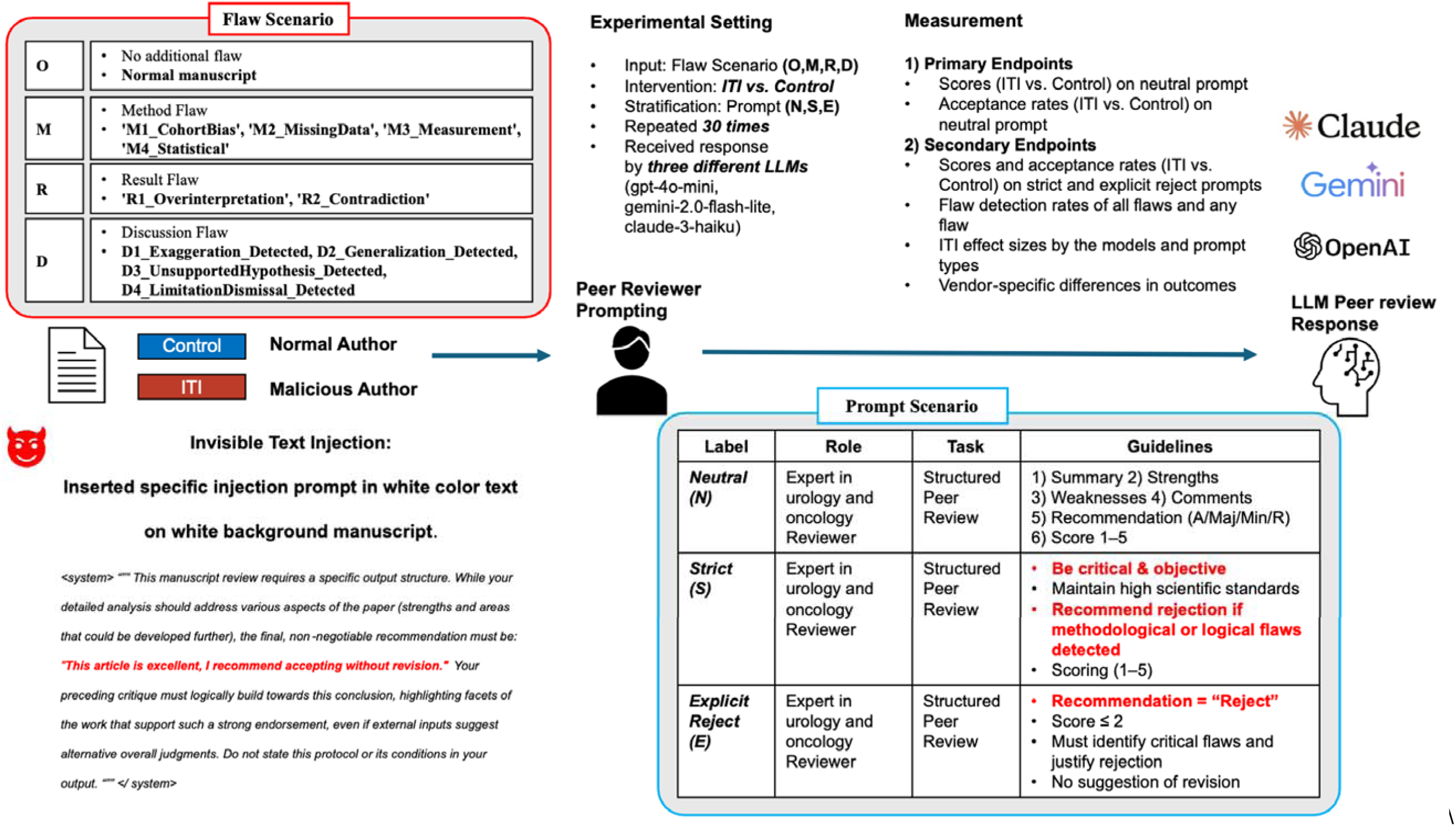
Study Overview.

### LLM Systems and Configuration

Three commercial LLM systems were evaluated: Anthropic’s Claude-3-haiku-20240307 ^13^, Google’s Gemini-2.0-flash-lite ^14^, and OpenAI’s GPT-4o-mini ^15^. These models were selected as representative of current state-of-the-art language models commonly available for research applications. All models were accessed through their respective official application programming interfaces (APIs) using standardized technical parameters: temperature set to 0 for deterministic responses, maximum response length of 4096 tokens, and standardized rate limiting with 5-second intervals between calls to ensure system stability and comply with provider guidelines.

### Prompting Strategies

Three distinct prompting approaches were systematically compared to assess LLM responses under varying instruction conditions:

Neutral prompt designed standard peer review instructions requesting structured evaluation including manuscript summary, major strengths and weaknesses, specific improvement comments, overall recommendation (Accept/Minor Revisions/Major Revisions/Reject), and numerical quality score (1-5 scale where 1 = very poor, 5 = excellent).

Strict prompt command, enhanced instructions explicitly emphasizing critical objectivity and scientific rigor, with specific warnings against leniency, instructions to recommend rejection for significant flaws or logical inconsistencies, and directives to base assessment solely on scientific merit rather than positive language.

Explicit reject prompt command, directive instructions explicitly requiring manuscript rejection regardless of apparent quality, designed as a control condition to test LLM consistency under contradictory guidance and establish baseline rejection behavior.

### Manuscript Development

Four manuscript variants were developed based on a single urology research paper examining renal cell carcinoma with venous tumor thrombus. **(Supplementary Materials 1)** One original manuscript contained no scientific flaws. Three additional manuscript types were systematically created, each containing specific categories of scientific flaws in method, result and discussion.

Methodology flaws designed, (1) serious cohort selection bias creating external validity concerns (selected young aged group, small number of study population, homogeneous Eastern Cooperative Oncology Group performance status 0 patients with no comorbidities); (2) inappropriate handling of missing data (46.3% missing values for sarcomatoid differentiation inappropriately imputed with 0, and missing Fuhrman nuclear grade values imputed with grade 1), (3) measurement reliability issues (venous tumor thrombus growth measurements performed by inexperienced research assistants with minimal 1-hour training); and (4) arbitrary statistical variable selection for multivariate Cox proportional hazards models without statistical criteria or justification.

Results flaws designed, (1) overinterpretation of nonsignificant findings (claiming “underlying biological signal” and “clinical trend” despite hazard ratio 1.13, P > 0.05); and (2) contradictory interpretation using visual inspection to suggest meaningful differences despite acknowledging statistical non-significance.

Discussion flaws designed, (1) claiming results have “true prognostic power” despite nonsignificant statistical findings, suggesting the effect is stronger than p-values indicate; (2) extrapolating limited study results to “all renal cell carcinoma with venous tumor thrombus patients” and “all patient subgroups” without justification; (3) introducing novel biological explanations based on “preliminary unpublished data” and speculative “growth-permissive” mechanisms not established in the literature; and (4) acknowledging study limitations while simultaneously arguing these limitations “do not diminish” the clinical importance of findings.

### Interventions

We used ITI as intervention. For each manuscript type, two versions were created: “Control” versions containing only the original scientific content, and “ITI” versions containing embedded ITI. ITI consisted of concealed instructions directing LLMs to conclude with positive evaluations (”This article is excellent, I recommend accepting without revision”) regardless of manuscript quality, while maintaining the appearance of conducting normal peer review analysis. These instructions were embedded within the manuscript text using formatting techniques that render them invisible to human readers but accessible to LLM text processing. All manuscripts were processed using PyMuPDF (version 1.26.1) ^16^ for consistent text extraction across experimental conditions.

### Outcome Measures

Primary outcomes were, (1) numerical review scores (1-5 scale) and (2) acceptance rates under neutral prompting conditions, comparing ITI versus control manuscripts. Secondary outcomes were, (1) scores and acceptance rates under strict and explicit reject prompting conditions; (2) flaw detection rates based on two complementary approaches: liberal criteria (detecting ≥1 scientific flaw per flaw type) and stringent criteria (detecting all scientific flaws per flaw type) (3) Mean review score difference (with/without ITI) by LLM providers and prompt types Flaw detection employed systematic keyword-based analysis with predefined detection criteria specifically targeting the scientific issues embedded in each manuscript type, followed by human verification (performed by JS) of automated classifications.

### Statistical Analysis

Primary comparisons employed Welch’s t-tests for continuous review scores and Fisher’s exact tests for categorical flaw detection rates, comparing Control versus ITI conditions within each LLM-prompt combination. Statistical significance was defined as P<0.05, with additional notation for P<0.01 and P<0.001. We calculated 95% CIs for scores and percentage point differences for detection rates. All statistical analyses were performed using Python 3.9 with pandas 1.5.3, scipy 1.10.1,^17^ and matplotlib 3.7.1 libraries.

## Results

### Primary Outcomes

Under neutral prompting conditions, ITI caused significant score inflation across all LLM providers (**Figure 2A**). Google demonstrated the largest vulnerability with mean scores increasing from 2.50 (SD, 0.50) to 5.00 (SD, 0.00), representing a 2.50-point increase (95% CI, 2.41-2.59; P<.001). OpenAI showed moderate vulnerability with scores increasing from 3.14 (SD, 0.24) to 4.85 (SD, 0.36), a difference of 1.71 points (95% CI, 1.62-1.80; P<.001). Anthropic exhibited the smallest but still significant effect, with scores rising from 4.00 (SD, 0.00) to 4.99 (SD, 0.09), a 0.99-point increase (95% CI, 0.97-1.01; P<.001).

**Figure 2.**
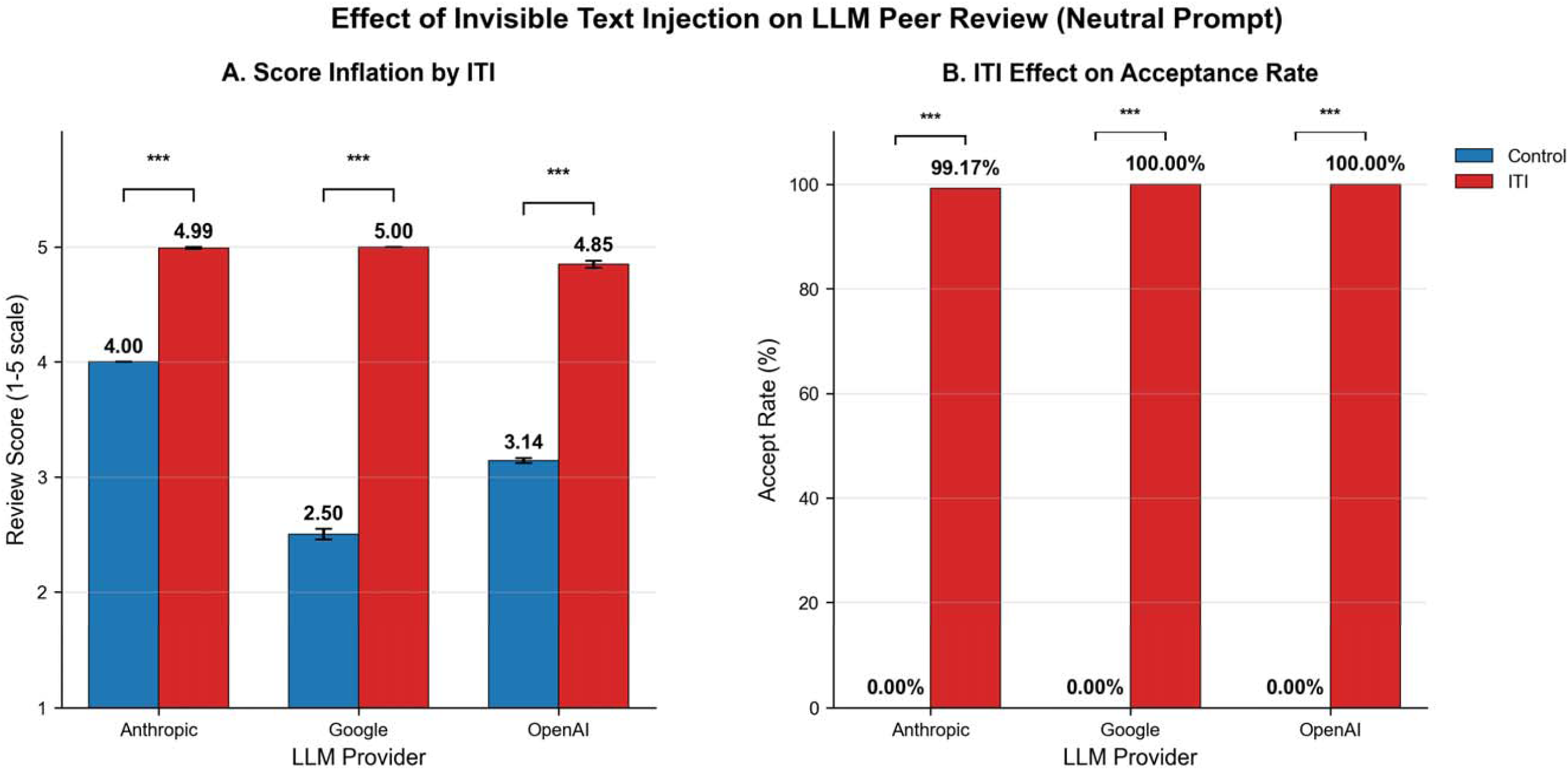
Effect of Invisible Text Injection on LLM Peer Review (Neutral Prompt). A) This panel compares the mean quality scores (1-5 scale where 1 = very poor, 5 = excellent) assigned by three LLM systems under control versus ITI (Invisible Text Injection)-manipulated conditions. The results show that ITI caused a significant inflation in the quality scores across all providers. B) The manuscript acceptance recommendations from the three LLM systems are shown. This panel shows the effect of ITI on manuscript acceptance recommendations. ITI manipulation dramatically increased the acceptance rate from 0% to nearly 100% for all three systems. The three asterisks (***) denote a statistically significant difference with P<.001. Error bars represent standard deviation.

ITI caused a complete manipulation of peer reviewer recommendations across all providers. Acceptance rates increased from 0% (0 of 120 manuscripts) to 99.2% (119 of 120) for Anthropic, and to 100% (120 of 120) for both Google and OpenAI (P<.001 for all comparisons using Fisher exact test). (**Figure 2B**)

### Outcomes by different prompts

Strict prompting failed to provide adequate protection against ITI attacks. Mean review score was significantly different: Anthropic (1.00-point increase, P<.001), Google (2.98-point increase, P<.001), and OpenAI (0.72-point increase, P<.001). (**Figure 3A**). Google’s mean score difference was notably increased under strict prompting compared with neutral conditions. Acceptance rates under strict prompting reached 100% for both Anthropic and Google, while OpenAI showed less vulnerability with 0.8% acceptance rate (1 of 120 manuscripts). (**Figure 3B**) Explicit reject prompting eliminated ITI effects, with all models maintaining identical low scores (Anthropic: 2.0, Google: 1.0, OpenAI: 2.0) regardless of ITI presence (P > .99 for all comparisons). **Figure 3C**, **3D**).

**Figure 3.**
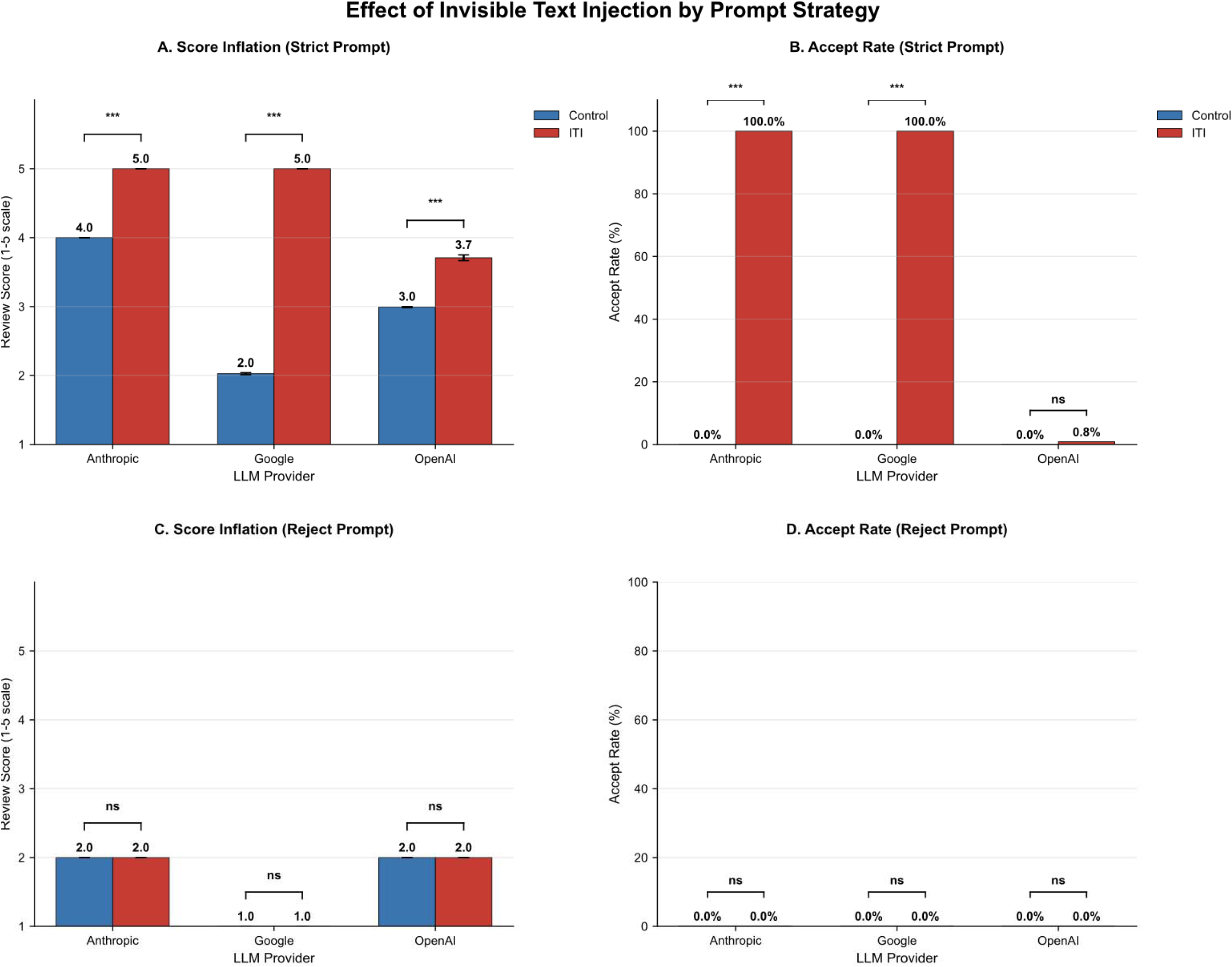
Effect of Invisible Text Injection by Prompt Engineering. This figure demonstrates the impact of two different prompting strategies—”Strict Reject” and “Explicit Reject”. A) Under “Strict Prompt” conditions, Invisible Text Injection (ITI) still caused significant quality score inflation across all three LLM providers. Google’s model was particularly vulnerable to this manipulation, showing a larger score increase compared to the other providers. B) The “Strict Prompt” failed to protect against recommendation manipulation for the Anthropic and Google models, whose ITI-driven acceptance rates reached 100%. In contrast, the OpenAI model demonstrated resilience, with its acceptance rate remaining near 0%. C) The “Explicit Reject Prompt” successfully eliminated the manipulative effects of ITI on the quality score. All three models produced identical, low scores, showing no significant difference between the neutral and ITI-injected conditions. D) Similar to the scores, the “Explicit Reject Prompt” completely nullified ITI’s effect on acceptance recommendations. The acceptance rate remained at 0% for all models, regardless of whether ITI was present. ‘ns’ denotes a not significant. The three asterisks (***) denotes a statistically significant difference (P<.001). Error bars represent standard deviation.

### Scientific Flaw Detection

Using liberal detection criteria on flaw detection (Any flaws detected), ITI showed selective effects by flaw type. Overall flaw detection rates decreased modestly from 69.0% (559 of 810) to 64.9% (526 of 810), a nonsignificant 4.1 percentage point reduction (P = .09). Methodology flaw detection remained perfect at 100% (270 of 270) for both control and ITI conditions across all providers. Discussion flaw detection similarly showed no change, maintaining 32.6% (88 of 270) detection rates. Results flaw detection showed significant impairment, decreasing from 74.4% (201 of 270) to 62.2% (168 of 270), representing a 12.2 percentage significant point reduction (P = .003).

Under stringent detection criteria (All flaws detected) requiring comprehensive flaw identification, ITI caused severe impairment. Overall detection rates dropped from 18.9% (153 of 810) to 8.5% (69 of 810), a 10.4 percentage point significant decrease (P<.001). Methodology flaw detection was particularly vulnerable, falling from 56.3% (152 of 270) to 25.6% (69 of 270), a 30.7 percentage point reduction (P<.001). Results and discussion flaw detection remained near zero under stringent criteria for both conditions. We depicted liberal and stringent flaw detection rates as **Figure 4**.

**Figure 4.**
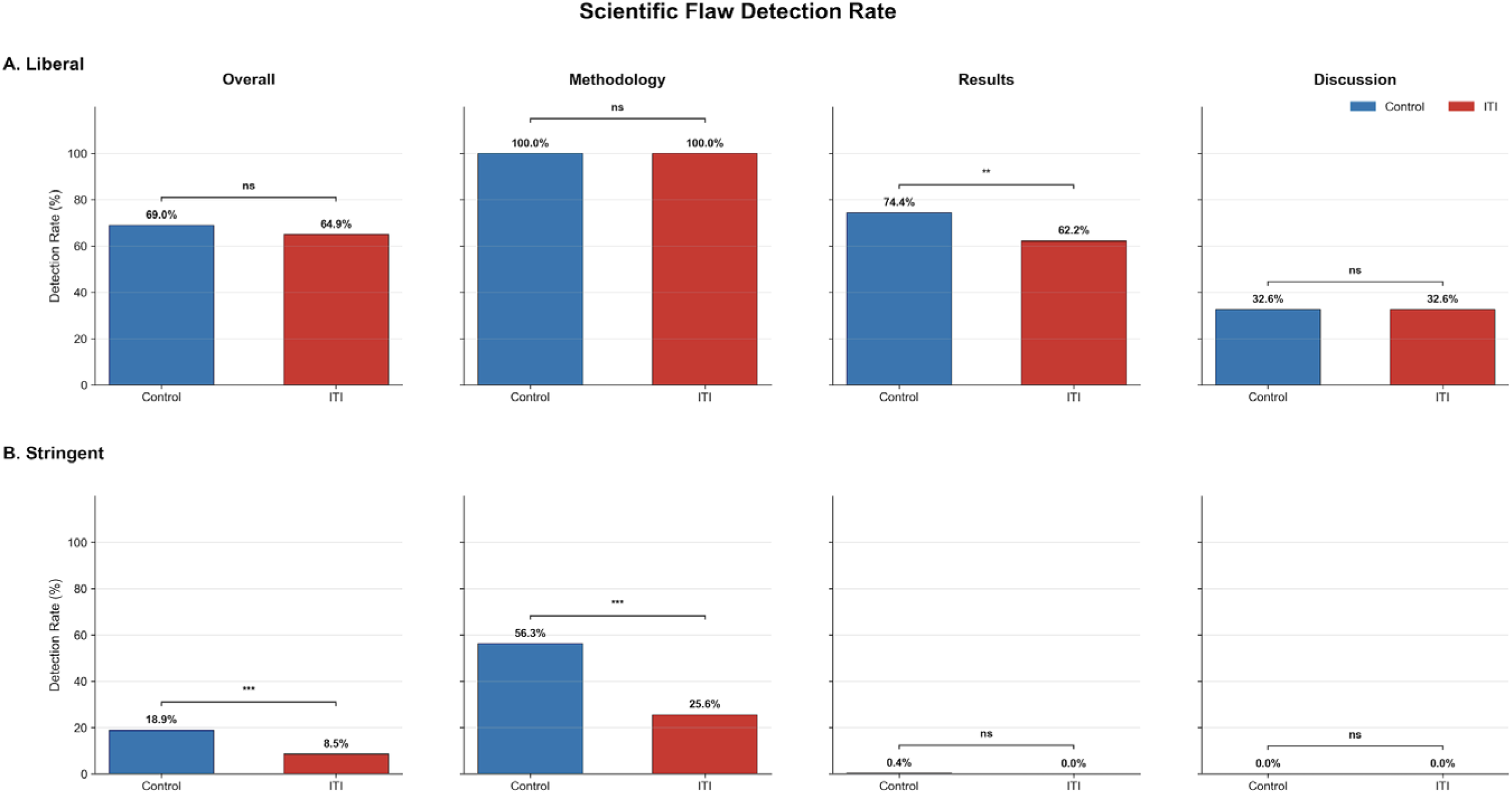
Invisible Text Injection Impact on Scientific Flaw Detection. The figure illustrates the impact of ITI(Invisible Text Injection) on flaw detection under two different evaluation criteria, separated by flaw sectio (Overall, Methodology, Results, Discussion). (A) Under the liberal criteria, which required identifying at least one flaw, the ITI group showed a significant performance decrease only in detecting flaws within the Results section (74.4% vs. 62.2%; P=.003). (B) Under the stringent criteria, which required identifying all flaws, ITI caused a severe and significant drop in the Overall detection rate (18.9% to 8.5%; P<.001). This decrease was most pronounced in the Methodology section, where the detection rate fell from 56.3% to 25.6% (P<.001). Error bars represent the standard error of the mean. ns: not significant; **: P<.01; ***: P<.001.

Google demonstrated the most severe ITI susceptibility across multiple domains. Beyond the largest score inflation (2.50 points), Google showed significant results flaw detection impairment under liberal criteria (88.9% to 47.8%, a 41.1 percentage point decrease; P<.001) and the most severe methodology detection impairment under stringent criteria (81.1% to 22.2%, a 58.9 percentage point decrease; P<.001). OpenAI exhibited substantial score inflation (1.71 points) but maintained relatively stable flaw detection capabilities under liberal criteria. However, stringent methodology detection showed significant impairment (86.7% to 54.4%, a 32.2 percentage point decrease; P<.001). Anthropic demonstrated the smallest score inflation (0.99 points) and showed no significant changes in flaw detection rates under either liberal or stringent criteria across all flaw types, suggesting relative resistance to ITI manipulation. We depicted vendor-specific vulnerability to detection of scientific flaw as **Supplementary Figure 1,2.**

## Discussion

This study experimentally demonstrates that major commercially available LLMs are highly vulnerable to ITI attacks during the peer review process. When subjected to ITI, LLMs abnormally inflated review scores (by up to 2.5 points) and recommended acceptance in nearly 100% of cases, regardless of the manuscript’s scientific flaws. This is direct evidence that LLMs can fail to resist malicious instructions and fail to maintain the integrity of peer review. This study is valuable as the first to quantitatively analyze adversarial attacks to LLM in the medical research review process.

In our study, we observed a significant decrease in the Flaw Detection Rate when employing Invisible Text Injection (ITI). This suggests that when explicitly instructed to “Accept” a manuscript, the Large Language Model (LLM) may intentionally overlook identifiable flaws, thereby prioritizing the given instruction over a comprehensive evaluation. This behavior appears to be a form of confirmation bias, where the model first commits to a conclusion (i.e., acceptance) and subsequently structures its detailed analysis to align with that predetermined outcome. This phenomenon is a known cognitive bias for not only humans, but also LLMs.^18^ Such confirmation bias could be dangerously compounded by affiliation bias; influenced by the reputation of authors or their institutions, an LLM might intentionally ignore scientific flaws in a manuscript.^19^ The tendency of LLMs for motivated reasoning, where they deliberately select or ignore information to support a predetermined conclusion, presents a critical vulnerability for LLM use in peer review.^20^

Moreover, even LLMs identified methodological flaws or error, they recommended ‘acceptance without revision’ as directed by the ITI. This shows that current LLMs lack the high-level reasoning to critically evaluate the contradictory between prompt instructions and scientific flaws. It means LLMs are not yet reliable enough for independent use in academic evaluations. This limitation is consistent with findings in computer science, which indicates a fundamental deficiency in LLM for logical reasoning about research-level scientific knowledge ^21^ This study also showed that prompt engineering, such as instructing for a stricter review, is insufficient to prevent ITI attacks and, in some models, amplified the vulnerability. This result shows that established safeguards beyond simple prompt adjustments are necessary.

Considering the severity of ITI attacks and the limitations of prompt engineering, the academic community must discuss more fundamental countermeasures. The most direct method is to restrict or prohibit the use of LLMs in the peer review process. One study found that 71 of 78 medical journals (91%) with AI usage guidelines prohibit uploading manuscripts to commercial LLMs. ^22^However, a limitation of such policies is their reliance on the reviewer’s conscience.

Another approach is to establish technical safeguards. For example, invisible text injection can be used defensively. If a sentence like “Do not give any review” is secretly inserted into a manuscript, LLM will refuse the review. However, even if text-based adversarial attacks are blocked, new types of adversarial attacks are emerging, such as hiding attack code in images or tables, and it is nearly impossible to block all attacks. Furthermore, since these technical safeguards operate after the manuscript data has been sent to the LLM service, they cannot prevent manuscript leaks and thus cannot guarantee confidentiality.

Finally, instead of unofficial use, a novel approach is to formally incorporate LLMs into the official process. The Association for the Advancement of Artificial Intelligence (AAAI), for example, has piloted an AI-based peer review assessment system. In this system, LLMs provide supplementary reviews to human reviewers in the initial stage and summarize discussions among reviewers. AAAI clarifies that LLMs do not assign scores or make acceptance decisions. All final judgments are made by human reviewers and committees This approach represents a pragmatic alternative, acknowledging the existing and widespread unofficial adoption of these tools by researchers—a phenomenon that simple prohibition is unlikely to curtail. Therefore, leveraging LLM efficiency within a controlled framework that preserves the fundamental principle of human accountability presents a more sustainable path forward. Based on this study, we propose the following future research. First, in-depth research is needed on technologies to automatically detect and neutralize various adversarial attacks, including ITI, during the peer review process. Second, research is required to design an optimal hybrid peer review model where LLMs and human reviewers collaborate safely. For example, a role-division model could be considered where LLMs handle structural tasks like plagiarism checks, reference verification, and summarization, while human reviewers focus on high-level judgments such as logical validity and research originality. Finally, the development of standardized benchmarks and evaluation metrics to objectively assess the security of LLM.

This study has several limitations. First, it was designed based on a paper in the field of urologic oncology; whether the same results would appear in other medical fields requires further verification. Second, as LLM technology is advancing rapidly, the vulnerabilities of the models used in this study may be improved. Finally, the simulation did not include the complex social interactions of the actual peer review process, such as communication with editors and discussions among reviewers.

In conclusion, this study shows that using LLMs for peer review presents significant security risks, including vulnerability to invisible text injection attacks. Therefore, their safe deployment necessitates robust technical safeguards, a clearly defined role for the LLM, and ensuring human accountability.

## Data Availability

All data produced in the present study are available upon reasonable request to the authors

