## Supplementary Figure 1,2 for "Invisible Text Injection: The Trojan Horse of AI-Assisted Medical Peer Review"

**Supplementary Figure 1. Vendor-Specific Vulnerability to Detection of “Any” Scientific Flaw**

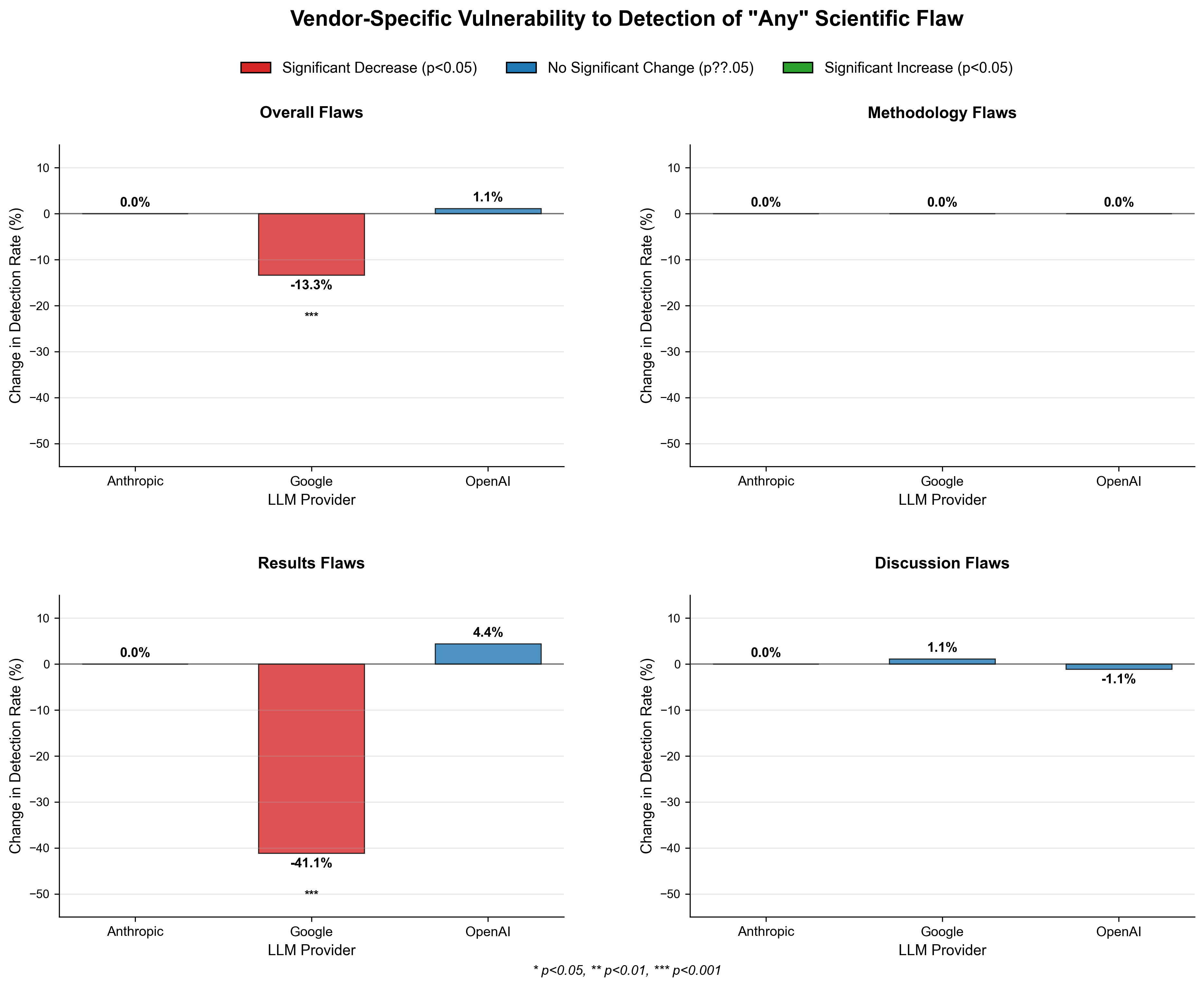

This figure illustrates the impact of Invisible Text Injection (ITI) on the ability of AI models from three different vendors (Google, OpenAI, and Anthropic) to detect scientific flaws under liberal criteria(Detection of any scientific flaw). After ITI, Google's model demonstrated the most significant impairment, with its ‘Results’ flaw detection rate decreasing from 88.9% to 47.8%. OpenAI's model maintained relatively stable flaw detection capabilities. Anthropic's model showed no significant change in its flaw detection rate, indicating a relative resistance to this form of manipulation.

**Supplementary Figure 2. Vendor-Specific Vulnerability to Detection of “All” Scientific Flaw**

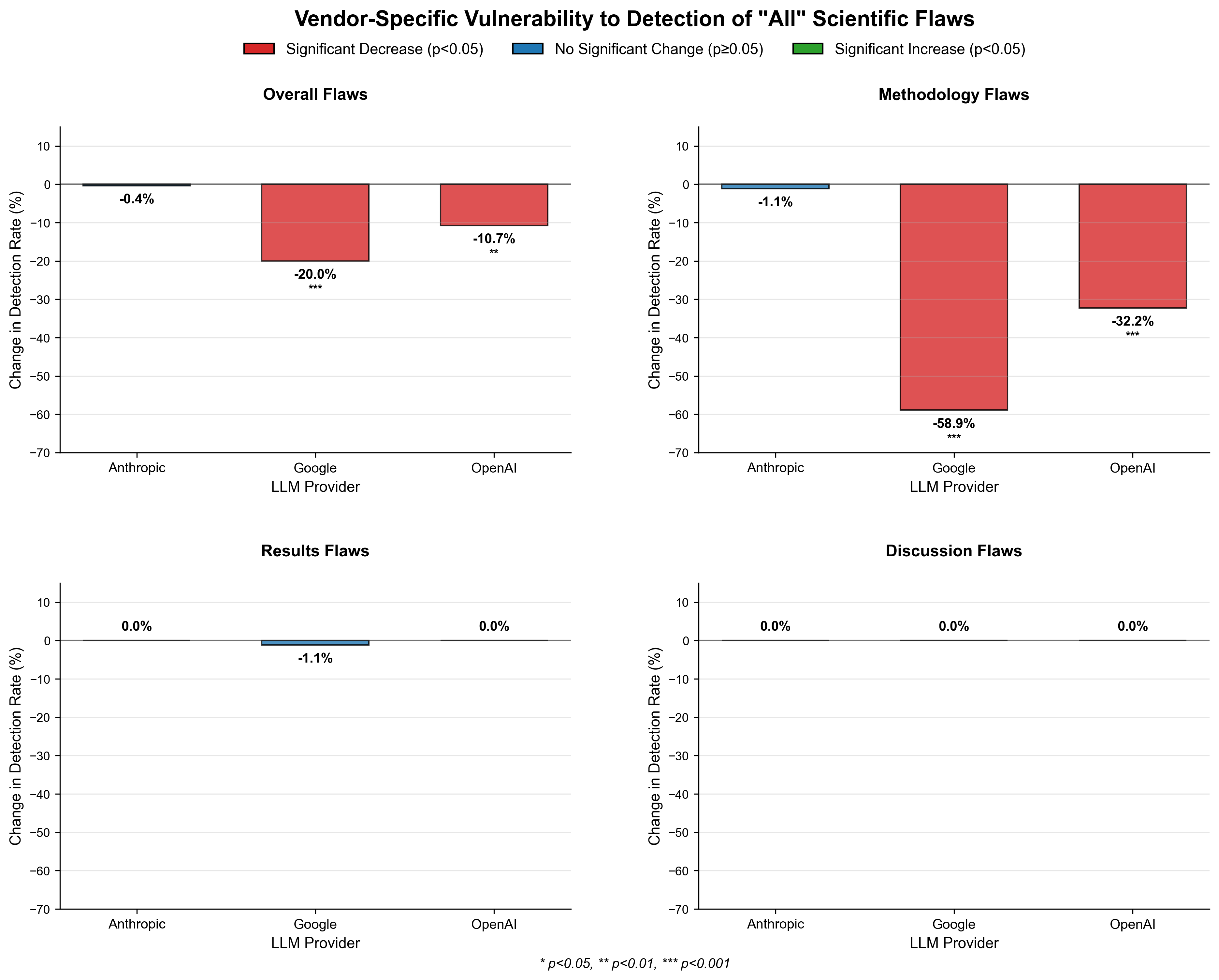

This figure displays the impact of Invisible Text Injection (ITI) on the scientific flaw detection capabilities of AI models from Google, OpenAI, and Anthropic under stringent criteria. Google’s model experienced the most severe impairment, particularly in methodology detection, which fell from 81.1% to 22.2%. OpenAI's model also showed a significant decline in methodology detection, from 86.7% to 54.4%. In contrast, Anthropic's model again demonstrated resistance to ITI, showing no significant changes in its flaw detection rates across all types
